# Factors Associated with Opioid Refills at 31-60 and 61-90 Days After Discharge from Spine Surgery: Potential Screening Marker for Transitional Pain Service

**DOI:** 10.1101/2025.07.02.25330755

**Authors:** Fang Ye, Lingyi Zhang, Praveen V. Mummaneni, Akash Shanmugam, Andrew Bishara, Sigurd H. Berven, Christopher R. Abrecht, Zhonghui Guan

## Abstract

**Background:** Although transitional pain service (TPS) has been proposed to manage patients at risk for persistent postsurgical opioid use, no standard criteria exist to identify high-risk patients for TPS management. Specifically, the 2-3 months following surgery are critical for the transition from short-term to persistent opioid use, yet little is known about opioid refills during this period.

**Methods:** This retrospective cohort study included 11,087 adult patients, regardless of opioid-naïve status, who underwent spine surgery at an academic medical center and were discharged between January 2017 and December 2023. Data were analyzed using multiple logistic and linear regressions and Fisher’s exact test.

**Results:** Of the patients, 25.4% and 14.8% received opioid refills at 31-60 and 61-90 days post-discharge, respectively. Among the independent risk factors, a refill at 31-60 days was the strongest predictor of a refill at 61-90 days (aOR 6.71, 95% CI 5.90-7.65), regardless of preoperative opioid use, cervical or lumbar procedures, or surgical service. Refill rates at 31-60 and 61-90 days were linearly correlated (p<0.0001, slope=0.73). A refill at 31-60 days predicted a refill at 61-90 days with a negative predictive value (NPV) of 94.3% and a positive predictive value of 41.5%, with consistently high NPVs across subgroups defined by preoperative opioid use, surgical procedure, or surgeon. A refill at 1-30 days; preoperative use of opioids, marijuana, and benzodiazepine; the first postoperative pain score recorded on hospital floor; and depression were all associated with increased odds of refills at both 31-60 and 61-90 days. In contrast, the total dose of discharge opioid prescriptions had minimal impact on refills.

**Conclusion:** A refill at 31-60 days after discharge may serve as a screening marker for identifying high-risk patients who could benefit from TPS management to mitigate further opioid use. Moreover, each refill prescription should be carefully managed to prevent subsequent refills.

## INTRODUCTION

Persistent postsurgical opioid use contributes to the ongoing opioid crisis^1–4^. To help prevent prolonged opioid use after surgery, transitional pain service (TPS) programs have been proposed to manage high-risk patients^1,5^. However, there is currently no standardized method to identify high-risk patients for TPS management, as each TPS program employs its own patient-selection criteria, making comparisons between different TPS programs and multi-center clinical trials difficult. Importantly, current patient-section criteria often fail to identify many patients who may benefit from TPS, as many so-called low-risk patients, such as opioid-naïve individuals who do not use opioids before surgery^6–9^ or those undergoing low-risk surgical procedures^2,10,11^, also develop persistent postsurgical opioid use.

Approximately 900,000 American adults undergo spine surgery annually^12^. Following cervical spine surgery, 6.3% of opioid-naive patients and 45.3% of non-opioid-naïve patients continued using opioids 12 months post-surgery^13^. After lumbar spine surgery, 8.6% of opioid-naive patients and 42.4% of non-opioid-naïve patients continued using opioids at 12 months^14^. By the third month after spine surgery, opioid use rates decreased substantially compared to the first and second months but were similar to those observed at 12 months post-surgery^13,14^.

While opioid use within the first month after surgery is considered immediate short-term use^15^, use beyond three months after surgery is classified as persistent opioid use^16–18^. Thus, the two to three months period after surgery represents a critical transitional phase from short-term to persistent opioid use. Although previous studies have examined risk factors associated with acute opioid use within the first postsurgical month^15^ and persistent opioid use beyond three months^10,11,18–23^, little is known about opioid use during the transitional two-to three-month postsurgical period for any type of surgical procedure, including spine surgery.

This study included 11,087 adult patients, regardless of opioid-naïve status, who underwent spine surgery, a procedure associated with high rates of persistent post-surgical opioid use^13,14^. The study aimed to identify independent risk factors associated with opioid refills at 31-60 days and 61-90 days after discharge and to develop a simple screening marker to detect high-risk patients likely to require further opioid refills following spine surgery.

## METHODS

### Study Design and Data Source

This retrospective observational cohort study was approved by the University of California San Francisco (UCSF) Institutional Review Board, which waived the requirement for patient consent to access electronic health record data.

#### Study Cohort

The cohort included 11,087 adult patients, regardless of opioid-naïve status, who underwent spine surgery performed by orthopedic spine and neuro-spine services at the UCSF Medical Center. Patients had postsurgical hospital stays of at least 1 day and were discharged between January 2017 and December 2023, a period following the release of the 2016 Centers for Disease Control and Prevention (CDC) guidelines for opioid prescriptions^24^.

### Outcome and Variables

The primary outcome was opioid refills prescribed at 31-60 and 61-90 days after hospital discharge. The study did not extend beyond 90 days, as many patients obtained opioid refills from their primary care physicians outside of the UCSF system after three months post-surgery, resulting in incomplete refill records beyond this period. Variables include preoperative opioid (within 6 months before surgery), marijuana, and benzodiazepine usage, first postsurgical pain level on hospital floor, last pain level before discharge, intra-operative oral morphine equivalents (OME), inpatient first daily OME after surgery, inpatient last daily OME before discharge, total OME of prescribed opioids at discharge, postsurgical usage of non-opioid analgesics (acetaminophen, non-steroidal anti-inflammatory drugs [NSAIDs], and gabapentinoids), opioid refill prescriptions at 1-30 days after discharge, type of the procedures (elective procedure, laminectomy, tumor resection, spine fusion, cervical procedures, lumbar procedures), surgical procedures (anterior cervical discectomy and fusion, anterior lumbar spine fusion 2-4 segments, cervical laminoplasty, endoscopic posterior spine fusion 1-2 segments, lateral lumbar interbody fusion, laminectomy for spinal cord tumor resection, lumbar decompression, posterior cervical spine fusion 4-7 segments, posterior lumbar spine fusion 1-3 segments, posterior lumbar spine fusion 4-6 segments, posterior lumbar fusion 7-12 segments, posterior spine fusion ≥13 segments, and posterior spine fusion with resection of spine tumor), neuro-spine vs ortho-spine service, procedure time in the operating room, return to operating room before discharge, postsurgical hospital length of stay (LOS), discharge disposition, psychiatric comorbidities (depression and anxiety), body mass index (BMI), age, sex, race, and ethnicity.

All opioid doses were converted to OME^25^. Similar to previous studies^26–29^, opioid daily dose over-prescription was defined as a maximum prescribed daily dose exceeding the patient’s last inpatient daily opioid consumption before discharge by at least 7.5 OME, while under-prescription was defined as a maximum prescribed daily dose of at least 7.5 OME below the patient’s last inpatient daily opioid consumption before discharge. 7.5 MME was set as the threshold because it corresponds to the dose of a 5 mg Oxycodone pill, the opioid medication most commonly prescribed opioid after surgery^30^, making it a clinically practical range for aligning daily doses.

### Statistical Analyses

Continuous variables, determined to be abnormally distributed by the D’Agostino and Pearson tests, were presented as medians with interquartile ranges (IQR). Binary variables were presented as percentages and analyzed using simple linear regression. Negative and positive predictive values were assessed using two-sided Fisher’s exact test. Multiple logistic regression was performed using the area under the ROC curve, a classification table, and predicted probability for each subject as classification and prediction methods, and the results were presented as adjusted odds ratios (aOR) and 95% confidence intervals (CI). All analyses were conducted using GraphPad Prism 10.3.1, with the statistical significance set at an α value of 0.05. A data analysis and statistical plan written and filed with institutional review board before data were accessed.

## RESULTS

### Demographic Data

As shown in Table 1, among the cohort of 11,087 patients with a median age of 64 years (IQR 54-72), 5,744 (51.8%) self-identified as female, 8,075 (72.8%) as white, 883 (8.0%) as Asian, 632 (5.7%) as black, 163 (1.5%) as Native American or Alaska Natives, 59 (0.5%) as Native Hawaiian or other Pacific Islanders, and 1,151 (10.4%) as Hispanic or Latino. Additionally, 6,243 (56.3%), 2,115 (19.1%), and 2,071 (18.7%) patients had preoperative use of opioids (within six months before surgery), marijuana, and benzodiazepines, respectively. During postoperative hospitalization, 10,386 (93.7%) patients received acetaminophen, 399 (3.6%) received NSAIDs, and 9974 (90.0%) received gabapentinoids (gabapentin or pregabalin).

**Table 1.**
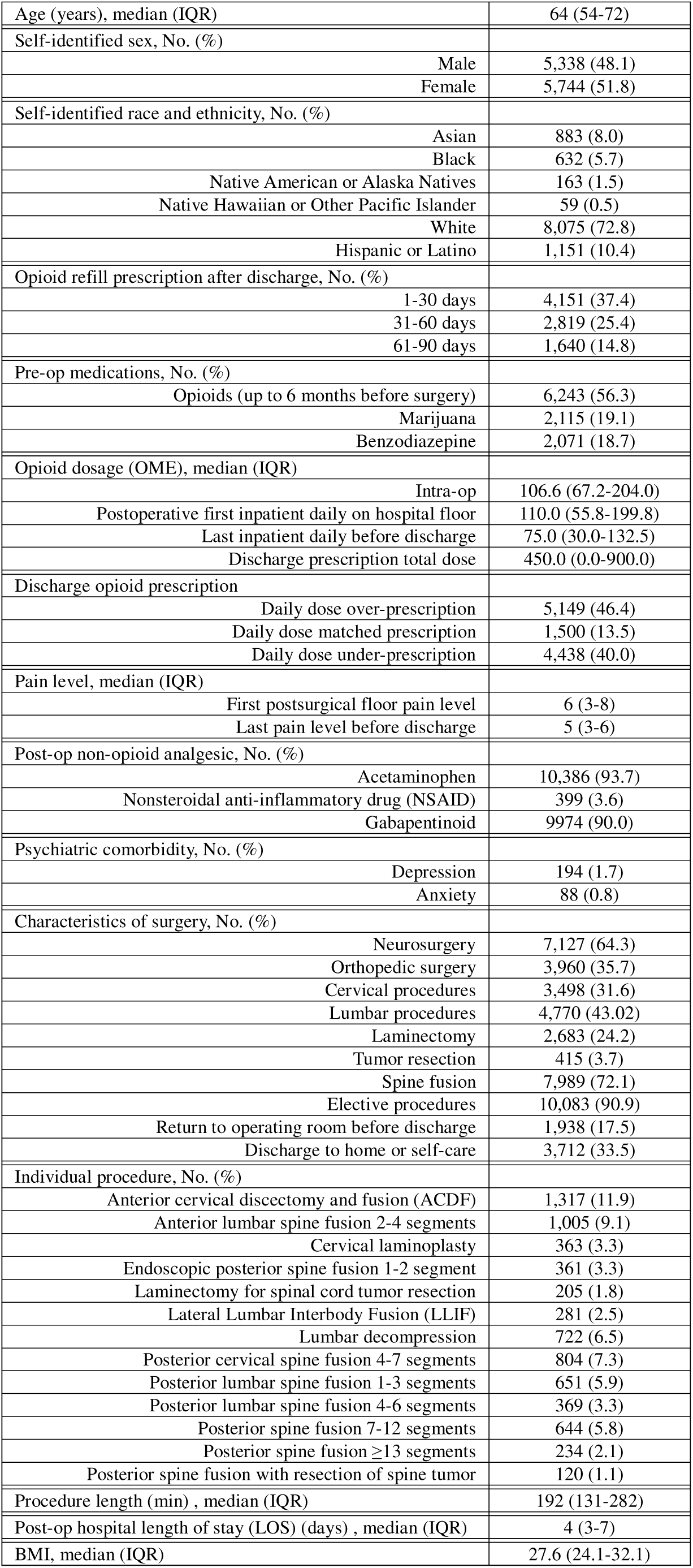
Demographic data of 11,087 spine surgical patients.

A total of 4,151 (37.4%), 2,819 (25.4%), and 1,640 (14.8%) patients received opioid refills at 1-30, 31-60, and 61-90 days after discharge, respectively. The median first pain level recorded on the hospital floor and last pain level recorded before discharge were 6 (IQR 3-8) and 5 (IQR 3-6) on a 0-10 scale, respectively. The median OMEs for intra-operative use, the first inpatient postsurgical daily consumption on the hospital floor, the last inpatient daily consumption before discharge, and the total dose of discharge opioid prescriptions were 106.6 (IQR 67.2-204.0), 110.0 (IQR 55.8-199.8), 75.0 (IQR 30.0-132.5) and 450 (IQR 0.0-900.0), respectively. A total of 5,149 (46.6%) patients were overprescribed opioid daily doses at discharge, while 4,438 (40.0) were under-prescribed.

### Previous Opioids Refills are Strongly Associated with Subsequent Opioid Refills

Our multiple logistic regression analyses (Table 2) revealed that, when controlling for other factors, an opioid refill at 31-60 days after discharge was the strongest risk factor associated with an increased odds of opioid refill at 61-90 days (aOR 6.71, 95% CI 5.90-7.65). Additionally, an opioid refill at 1-30 days was strongly associated with increased odds of refills at both 61-90 days (aOR 3.16, 95% CI 2.76-3.62) and 31-60 days (aOR 4.04, 95% CI 3.66-4.46) after discharge.

**Table 2.**
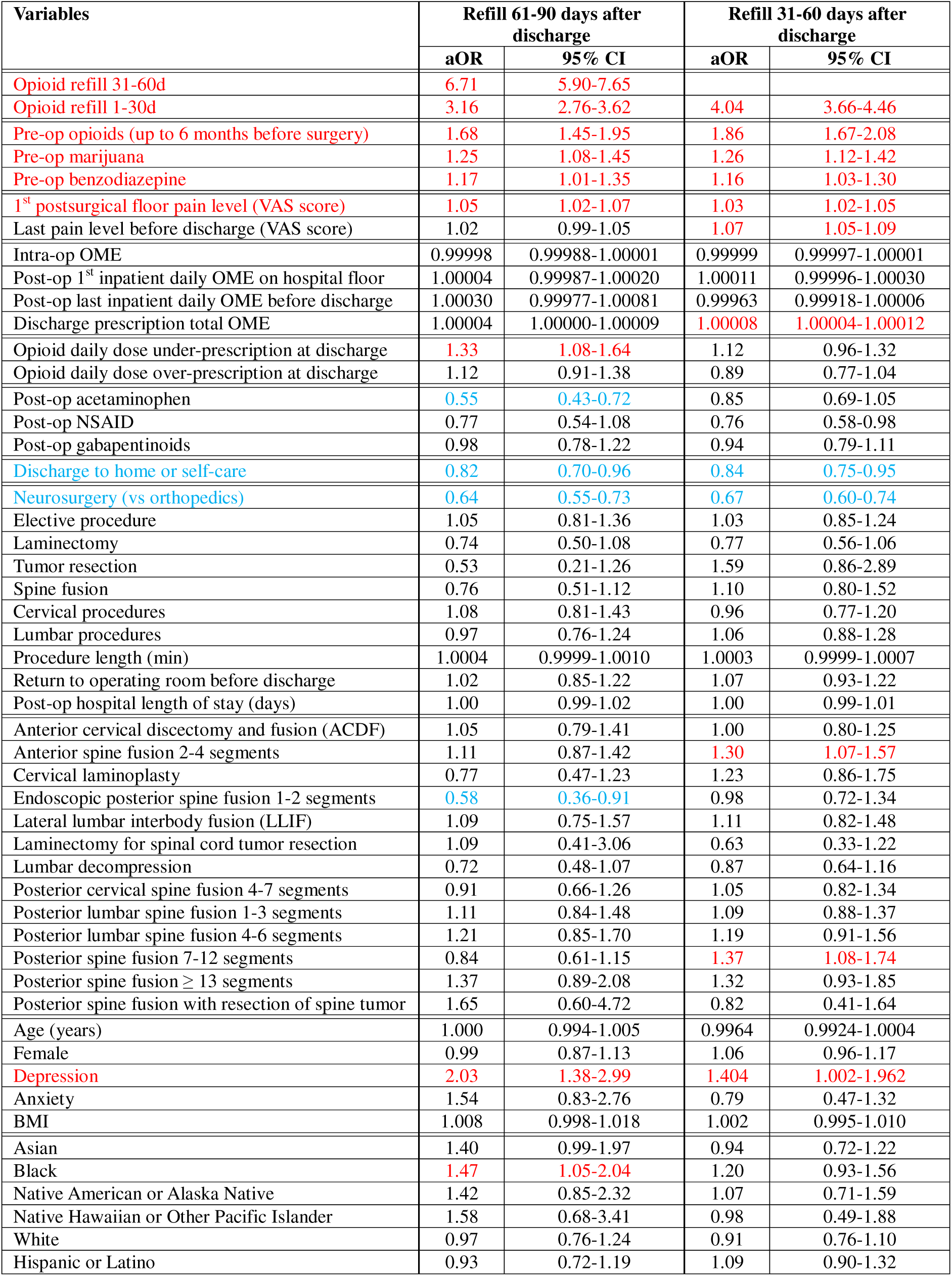
Independent risk factors associated with opioid refill prescriptions 61-90 and 31-60 days after discharge from spine surgeries. aOR: adjusted odds ratio; CI: confidence interval.

### Other Postoperative Factors Associated with Opioid Refills at 31-60 and 61-90 Days After Discharge

Multiple logistic regression analyses also revealed that when other factors were controlled, postsurgical acetaminophen (aOR 0.55, 95% CI 0.43-0.72) was associated with reduced odds of opioid refill at 61-90 days after discharge. Discharge to home or self-care was associated with reduced odds of refills at 61-90 days (aOR 0.82, 95% CI 0.70-0.96) and 31-61 days (aOR 0.84, 95% CI 0.75-0.95) after discharge, respectively.

Additionally, higher first postsurgical pain levels recorded on the ward floor were associated with increased odds of opioid refill at 61-90 (aOR 1.05, 95% CI 1.02-1.07) and 31-60 days (aOR 1.03, 95% CI 1.02-1.05), whereas higher last pain levels recorded before discharge were associated with an increased odds of refill at 31-60 days (aOR 1.07, 95% CI 1.05-1.09). The intraoperative OME, the first postoperative inpatient daily OME on hospital floor, and the last inpatient daily OME before discharge were not associated with increased or decreased odds of opioid refills. However, a higher total dose of discharge opioid prescription was associated with an increased odd of opioid refill at 31-60 days (aOR 1.00008, 95% CI 1.00004-1.00012), but not at 61-90 days after discharge. Opioid daily dose under-prescription at discharge was associated with an increased odds of refill at 61-90 days (aOR 1.33, 95% CI 1.08-1.64), while opioid daily dose over-prescription was not associated with opioid refills.

### Preoperative Factors Associated with Opioid Refills at 31-60 and 61-90 Days After Discharge

Preoperative opioid use within 6 months before surgery was associated with increased odds of opioid refills at 61-90 days (aOR 1.68, 95% CI 1.45-1.95) and 31-60 days (aOR 1.86, 95% CI 1.67-2.08) after discharge. Preoperative use of marijuana (aOR 1.25, 95% CI 1.08-1.45) and benzodiazepine (aOR 1.17, 95% CI 1.01-1.35), as well as depression (aOR 2.03, 95% CI 1.38-2.99), were also associated with an increased odds of refills at 61-90 days after discharge. Similarly, preoperative use of marijuana (aOR 1.26, 95% CI 1.12-1.42) and benzodiazepine (aOR 1.16, 95% CI 1.03-1.30), and depression (aOR 1.404, 95% CI 1.002-1.962), were also associated with increased odds of refills at 31-60 days after discharge as well. Age, sex, race, ethnicity, anxiety, and BMI were not associated with opioid refills, except that being black was associated with an increased odds of refill at 61-90 days (aOR 1.47, 95% CI 1.05-2.04).

### Association between Surgical Procedures and Opioid Refills at 31-60 and 61-90 Days After Discharge

Compared to orthopedic spine surgery, neuro-spine surgery was associated with decreased odds of opioid refills at 61-90 (aOR 0.64, 95% CI 0.55-0.73) and 31-60 days (aOR 0.67, 95% CI 0.60-0.74) when other factors were controlled. Overall, specific spine surgical procedures were generally not associated with increased or decreased odds of opioid refills. However, endoscopic posterior spine fusion of 1-2 segments was associated with a decreased odds of opioid refill at 61-90 days (aOR 0.58, 95% CI 0.36-0.91), while anterior spine fusion of 2-4 segments (aOR 1.30, 95% CI 1.07-1.57) and posterior spine fusion of 7-12 segments (aOR 1.37, 95% CI 1.08-1.74) were associated with an increased odds of opioid refill at 31-60 days.

### Trends in the Rates of Opioid Refill at 31-60 and 61-90 Days After Discharge

We analyzed the opioid refill rates at 31-60 and 61-90 days from 2017 to 2023 at six-month intervals. The refill rate at 31-60 days decreased significantly during this period (p = 0.003, slope =-0.39) (Figure 1a). When the refill rate at 31-60 days was plotted against that at 61-90 days, a linear correlation was observed (p < 0.0001, slope = 0.73) (Figure 1b).

**Figure 1.**
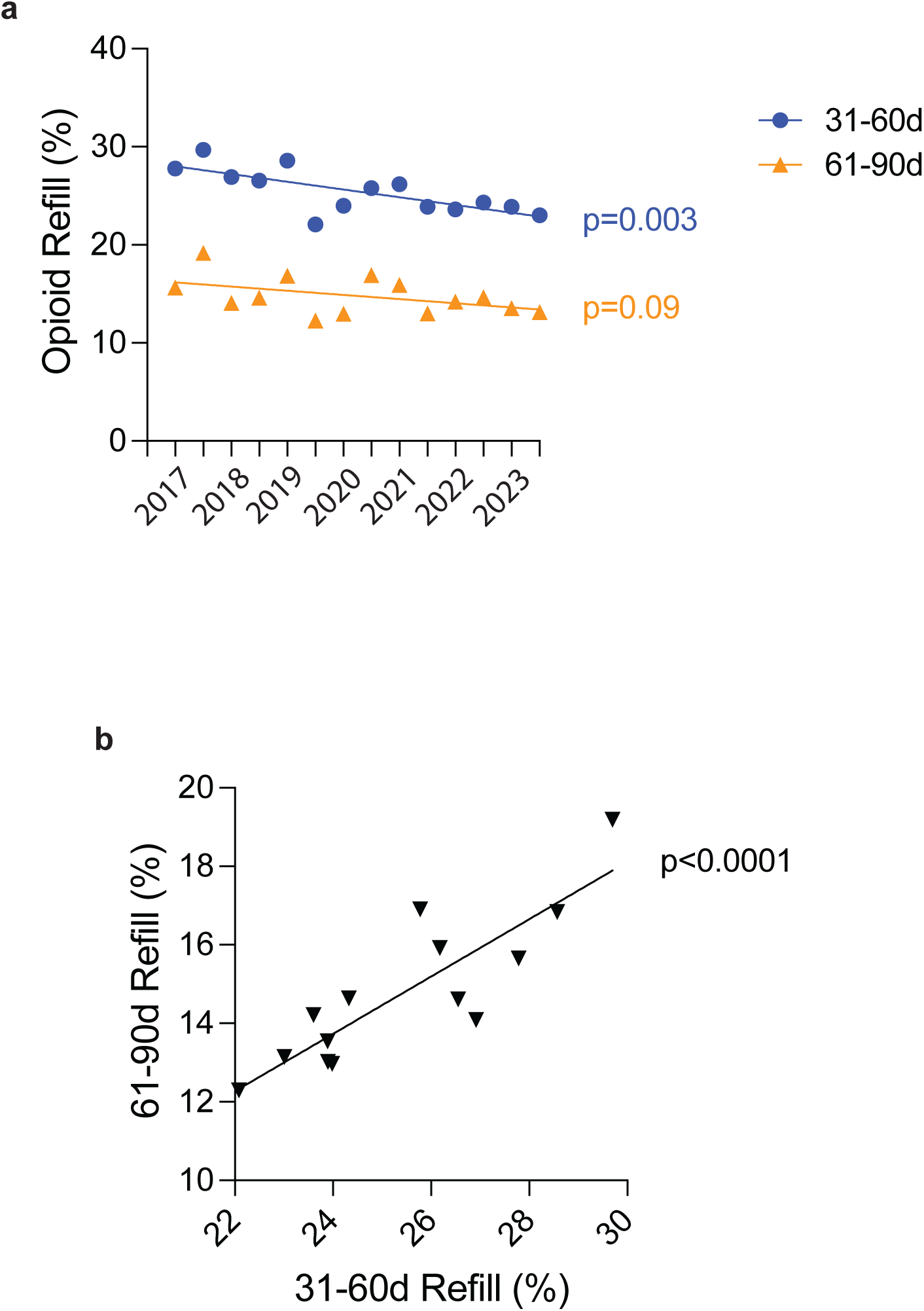
Rates of opioid refills at 31-60 days and 61-90 days after discharge from spine surgery. **a.** The rate of opioid refill at 31-60 days after discharge significantly decreased from 2017 to 2023. **b.** There is a linear relationship between the rates of opioid refills at 31-60 days and 61-90 days after discharge. Data were analyzed using linear regression.

### Strong Association Between Opioid Refills at 31-60 and 61-90 Days in Patients With and Without Pre-operative Opioids, Following Cervical and Lumbar Spine Surgeries, and by Neurosurgery and Orthopedic Services

Our multiple logistic regression analyses showed that, with other factors controlled, a refill at 31-60 was strongly associated with increased odds of refill at 61-90 days in patients with (aOR 6.28, 95% CI 5.40-7.31) and without (aOR 8.01, 95% CI 6.21-10.38) preoperative opioid use (Figure 2a), following cervical (aOR 6.06, 95% CI 4.74-7.76) and lumbar (aOR 7.60, 95% CI 6.27-9.24) spine surgeries (Figure 2b), as well as in patients receiving care from neurosurgery (aOR 6.24, 95% CI 5.23-7.46) and orthopedic (aOR 7.60, 95% CI 6.25-9.27) services (Figure 2c).

**Figure 2.**
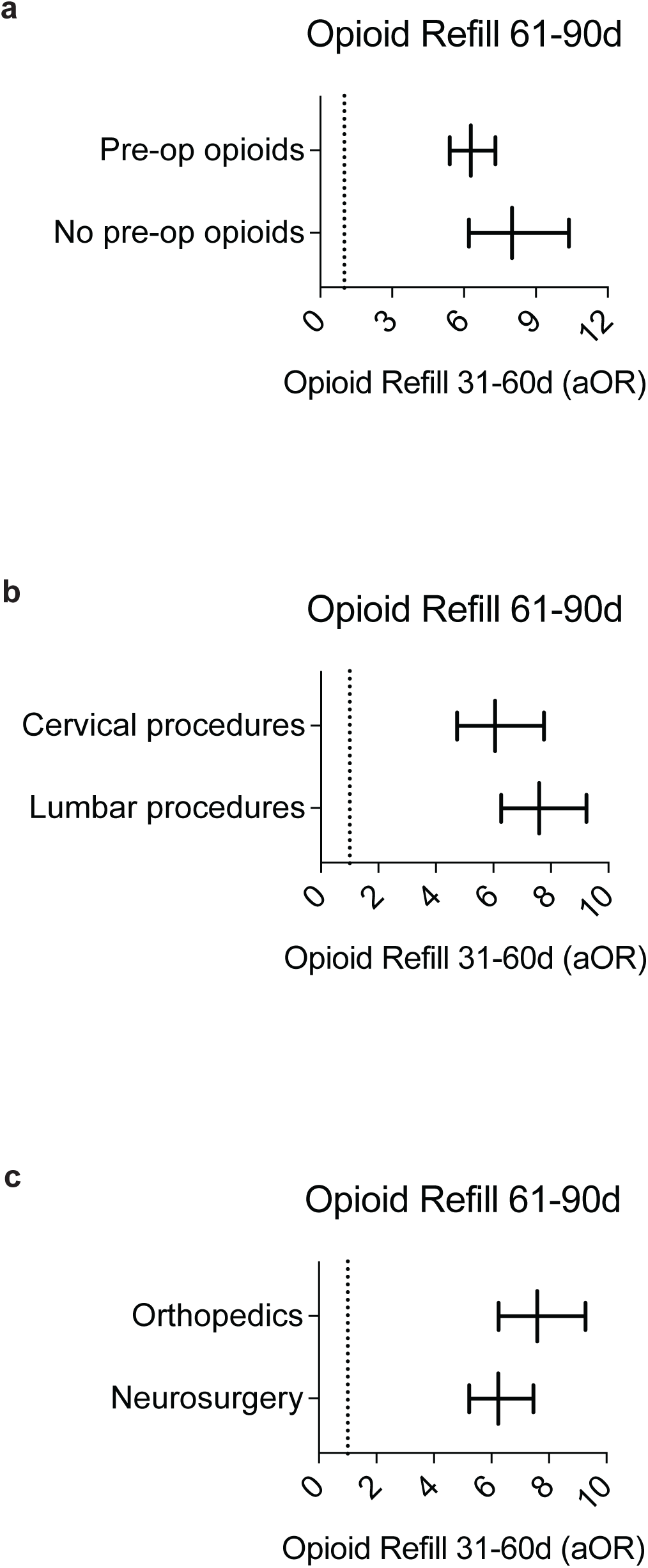
Strong association between opioid refills at 31-60 days and 61-90 days after discharge from spine surgery. With other variables controlled, opioid refills at 31-60 days after discharge were strongly and independently associated with opioid refills at 61-90 days among patients with or without pre-operative opioids (**a**), following cervical or lumbar procedures (**b**), and across neurosurgery or orthopedics (**c**). The other controlled variables are listed in the methods section. Data were analyzed using multiple logistic regression, with p<0.0001 for each analysis.

### Predicting Opioid Refill at 61-90 Days Based on Refill at 31-60 Days After Discharge

We explored the potential of using opioid refill at 31-60 days as a screening marker to predict subsequent refill at 61-90 days. This approach yielded a negative predictive value (NPV) of 94.3% (Figure 3a). Specifically, for patients with and without pre-operative opioid use, the NPVs were 92.0% and 96.7%, respectively. For patients undergoing different spine procedures, the NPVs exceeded 91% across all procedures (ranging from 91.7% in lateral lumbar interbody fusion to 98.9% in posterior spine fusion with tumor resection), except for posterior spine fusion of ≥13 segments, which had an NPV of 87.7%. Among the seven neurosurgeons and six orthopedic surgeons who performed more than 100 procedures during the study period, the NPVs ranged from 90.6% to 96.0% (Figure 3a). The positive predictive values (PPVs) for the prediction were 41.5% for the entire cohort, and 45.0% and 32.2% for patients with and without pre-operative opioid use, respectively. For patients undergoing different spine procedures, the PPVs ranged from 23.5% to 50.0%. Among the 13 surgeons, the PPVs ranged from 26.4% to 59.4% (Figure 3b).

**Figure 3.**
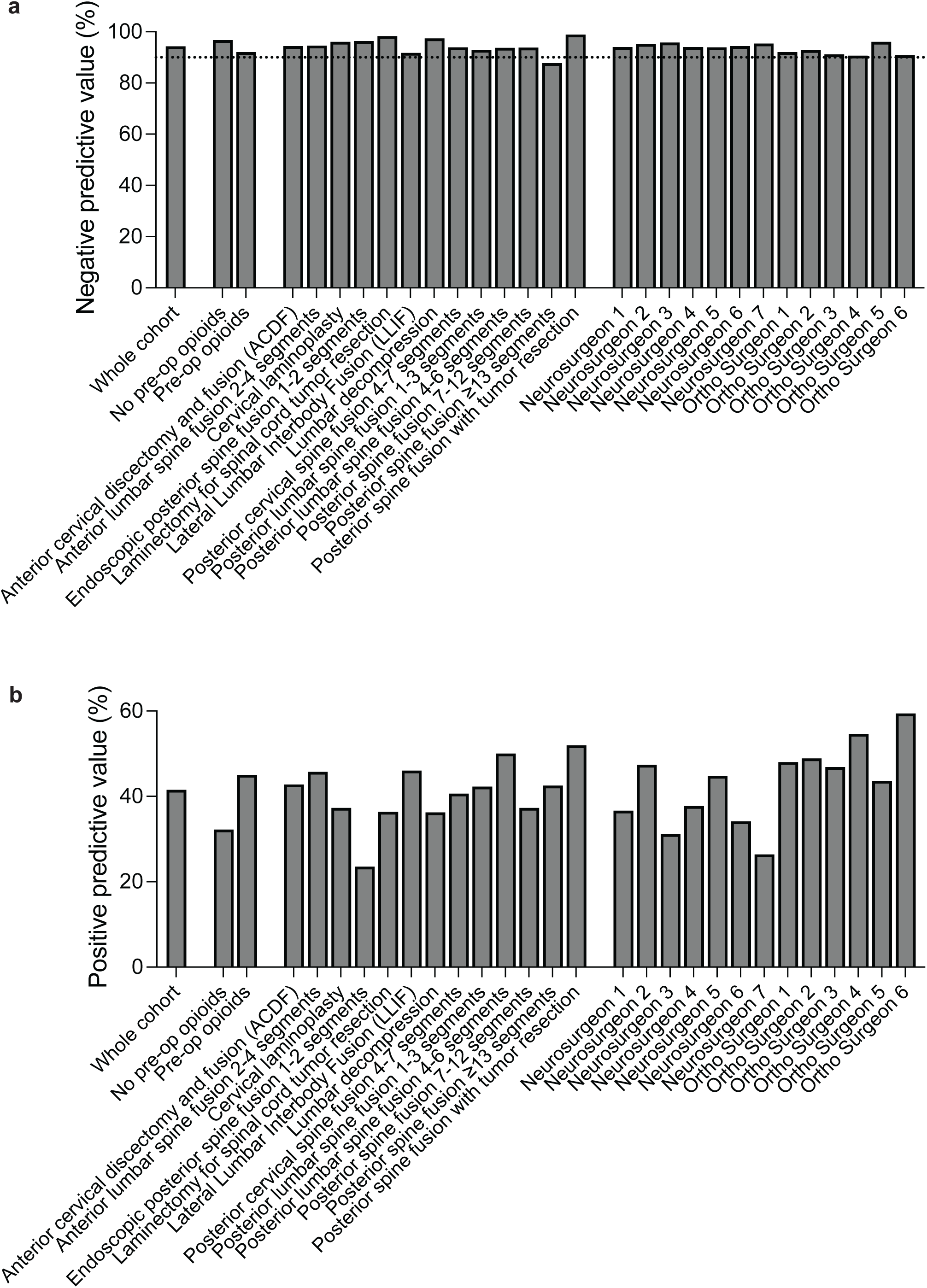
Using opioid refill at 31-60 days after discharge to predict a refill at 61-90 days. Negative (**a**) and positive (**b**) predictive values are presented for the whole cohort, with or without preoperative opioids, across different spine procedures, and by different surgeons. Data were analyzed using two-sided Fisher’s exact test, with p<0.0001 for each analysis.

Opioid refill at 1-30 days after discharge can also be used to predict refill at 61-90 days (Supplementary Figure 1) or refill at 31-60 days (Supplementary Figure 2); however, it provides much lower PPVs (for 61-90 days) and NPVs (for 31-60 days).

## DISCUSSION

To prevent persistent postsurgical opioid use beyond three months after surgery, it is crucial to implement interventions for high-risk patients before persistent use becomes established. However, little is known about opioid use during the transitional period of 2-3 months postoperatively. Moreover, although TPS has been proposed to prevent persistent postsurgical opioid use, there is no standardized method to identify high-risk patients. In this study, we identified independent factors associated with opioid refills prescribed at 31-60 days and 61-90 days after discharge from spine surgery, a procedure with a high incidence of persistent postsurgical opioid use^13,14^. Among these factors, opioid refill at 31-60 days was the strongest predictor of refill at 61-90 days, and the refill rates refills at 31-60 days and 61-90 days were linearly correlated. Using opioid refill at 31-60 days to predict refill at 61-90 days yielded an NPV of 94.3% and a PPV of 41.5%. Notably, the NPVs were consistently high regardless of preoperative opioid use, procedure, or surgeon.

We used opioid refill prescriptions as a proxy for opioid use in this study as prescriptions serve as a gateway for patients to access opioids^31^. We found that the strongest independent predictors of opioid refills at 31-60 days and 61-90 days were prior opioid refills. After controlling for other factors, a refill at 31-60 days after discharge was associated with a more than six-fold increase in the odds of refill at 61-90 days. This strong association persisted regardless of preoperative opioid use, whether the procedure was cervical or lumbar spine surgery, or whether care was provided by orthopedic or neurosurgical services. Conversely, refill at 1-30 days was associated with a more than three-fold increase in the odds of refill at 61-90 days and a four-fold increase in the odds of refill at 31-60 days after discharge. These findings highlight the importance of carefully managing each postsurgical opioid refill prescription following spine surgery to prevent subsequent refills.

As prevention is widely regarded as the most effective strategy for managing chronic diseases^32^, it is likely also the best approach for addressing persistent postsurgical opioid use. Therefore, the transitional phase of two to three months after surgery may represent a crucial window for implementing interventions to prevent long-term opioid use. Given the strong association and linear correlation between refills at 31-60 days and 61-90 days after discharge, an opioid refill at 31-60 days may serve as a useful screening marker to identify patients at high risk for subsequent refills at 61-90 days. This marker demonstrated an NPV of 94.3% and a PPV of 41.5%. In other words, if a patient does not receive opioid refill at 31-60 days, there is a 94.3% likelihood they will not require a refill at 61-90 days. Conversely, if a patient does receive a refill at 31-60 days, there is a 41.5% chance that they will require a refill at 61-90 days.

Therefore, it may be advisable to refer patients who receive a refill at 31-60 days after discharge to a TPS for multimodal management to prevent further opioid use following spine surgery. Since the opioid refill rate in the third month after spine surgery closely mirrors that at 12 months^13,14^, TPS interventions aimed at reducing opioid refill in the third month may help reduce opioid use by 12 months. Notably, during our 7-year study period, 2,819 spine surgery patients received refills at 31-60 days after discharge (Table 1), averaging fewer than eight patients per week, well within the manageable capacity of a TPS. Although refills at 1-30 days may also serve as a screening marker for refills at 31-60 and 61-90 days, their NPVs and PPVs are considerably lower.

Since many post-operative factors are associated with opioid refills at 31-60 and 61-90 days after discharge, efforts to prevent prolonged opioid use should begin shortly after spine surgery. For example, postsurgical pain should be adequately managed. For every 1-point increase in the first pain score (on a scale of 0-10) recorded on the hospital floor, the odds of opioid refills at 31-60 and 61-90 days increased by 3% and 5%, respectively. Additionally, each 1-point increase in the last pain score recorded before discharge was associated with a 7% increase in the odds of refill at 31-60 days. Notably, postsurgical administration of acetaminophen was associated with a reduced risk of opioid refill at 61-90 days, suggesting that its use provides long-term benefit and should be encouraged after spine surgery to reduce postsurgical pain and lower the likelihood of prolonged opioid refills. However, after adjusting for other factors, intraoperative OME, the first postsurgical inpatient daily OME, and the last inpatient daily OME before discharge were not associated with increased or decreased risks of opioid refills.

Although substantial efforts have focused on reducing the total dose of discharge opioid prescriptions based on procedure-centered guidelines^33–35^, our study showed that the total dose of discharge opioid prescriptions was not associated with an increased or decreased risk of opioid refills at 61-90 days after discharge. While it was associated with an increased risk of refills at 31-60 days, the effect was clinically marginal, with each 1 OME increase corresponding to only a 0.008% increase in odds, or a 0.8% increase in odds for every 100 OME increase. In contrast, opioid daily dose under-prescription, defined as the maximum daily dose of discharge opioid prescription being at least 7.5 OME lower than the patient’s last inpatient daily opioid consumption before discharge, was associated with a 33% increase in the odds of opioid refills at 61-90 days. Since poorly controlled acute pain is one of the strongest predictors of chronic postsurgical pain^5^, patients whose maximum daily dose of discharge opioid is lower than their daily opioid requirements may experience undertreated acute pain, placing them at higher risk of long-term pain and prolonged opioid use after spine surgery.

Preoperatively, opioid use within six months before surgery was associated with a 68% and 86% increase in the odds of opioid refills at 61-90 days and 31-60 days after discharge, respectively. Preoperative use of marijuana and benzodiazepine, as well as a history of depression, were also associated with increased risk of refills during both time periods. However, given the substantial contribution of many postsurgical factors to opioid refills, identifying high-risk patients based solely on preoperative factors can be challenging.

As an observational study, our research had the inherent limitations of unmeasured confounding variables. For example, while we investigated opioid refill prescriptions, we could not capture the actual amount of opioids consumed by the patients. However, since opioid prescription serves as a gateway for the opioids to be potentially accessed by patients and released into the community, proper prescribing practices are crucial for effective postsurgical opioid management. Additionally, although our analyses accounted for the preoperative use of opioids, marijuana, and benzodiazepines, our dataset did not include information on the preoperative use of other substances with abuse potential.

Our results suggest that each postsurgical opioid refill prescription after spine surgery should be carefully managed to prevent subsequent opioid refills. Given the strong association and linear correlation between refill rates at 31-60 and 61-90 days, an opioid refill at 31-60 days may serve as a simple yet effective screening marker to identify high-risk patients who could benefit from TPS management to reduce further opioid use after spine surgery.

## Abbreviations and acronyms

aOR: adjusted odds ratio
BMI: body mass index
CDC: Centers for Disease Control and Prevention
CI: confidence interval
IQR: interquartile range
LOS: length of stay
NPV: negative predictive value
OME: oral morphine equivalent
PPV: positive predictive value
TPS: transitional pain service
UCSF: University of California San Francisco

## Data Availability

All data produced in the present study are available upon reasonable request to the authors

---

**Supplementary Figure 1.**
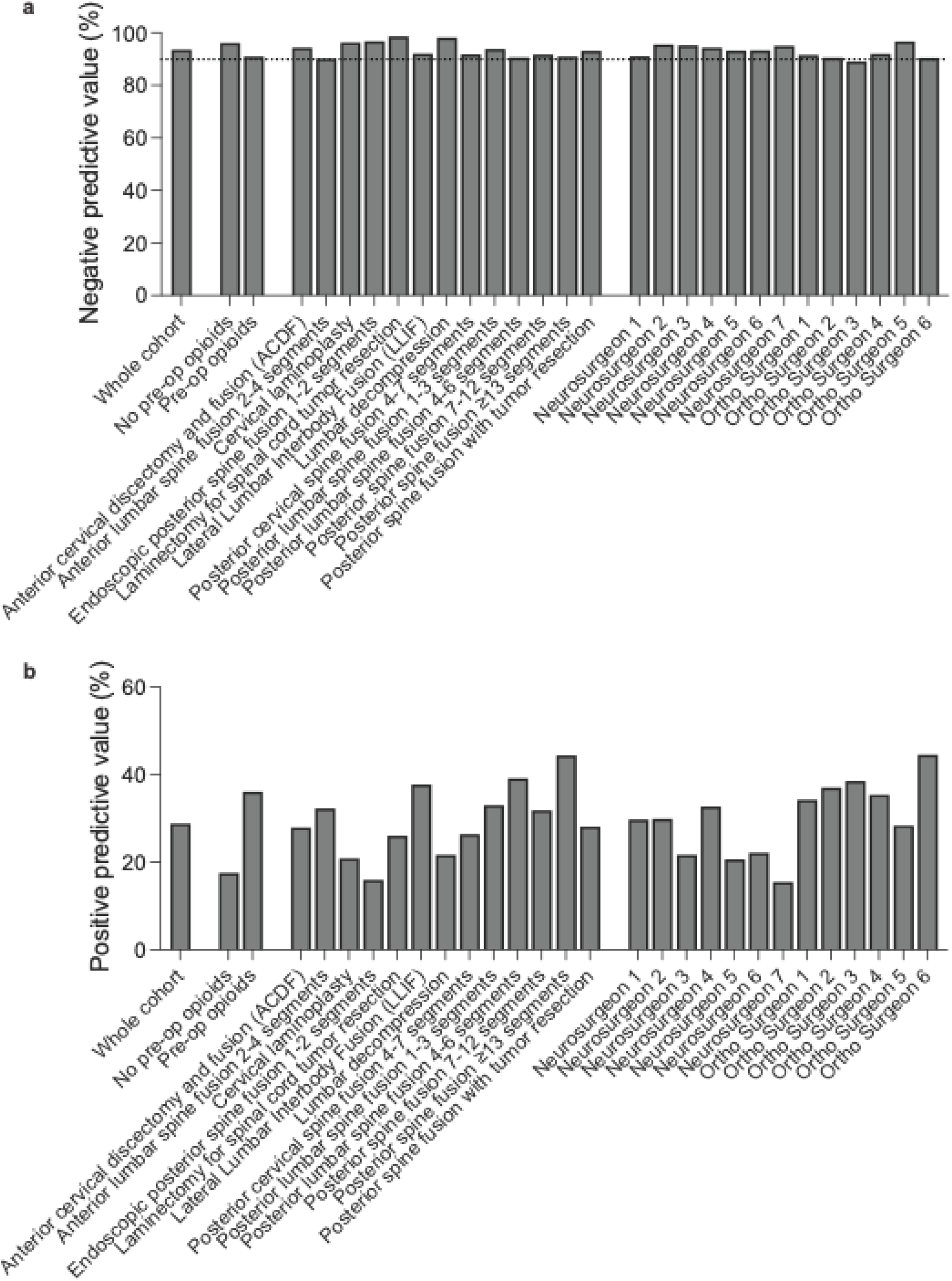
Using opioid refill at 1-30 days after discharge to predict a refill at 61-90 days. The negative (**a**) and positive (**b**) predictive values are presented for the whole cohort, with or without pre-operative opioids, across different procedures, and by different surgeons. Data were analyzed using two-sided Fisher’s exact test, with p<0.0001 for each analysis except posterior spine fusion with tumor resection and neurosurgeon 5, which have p<0.01.

**Supplementary Figure 2.**
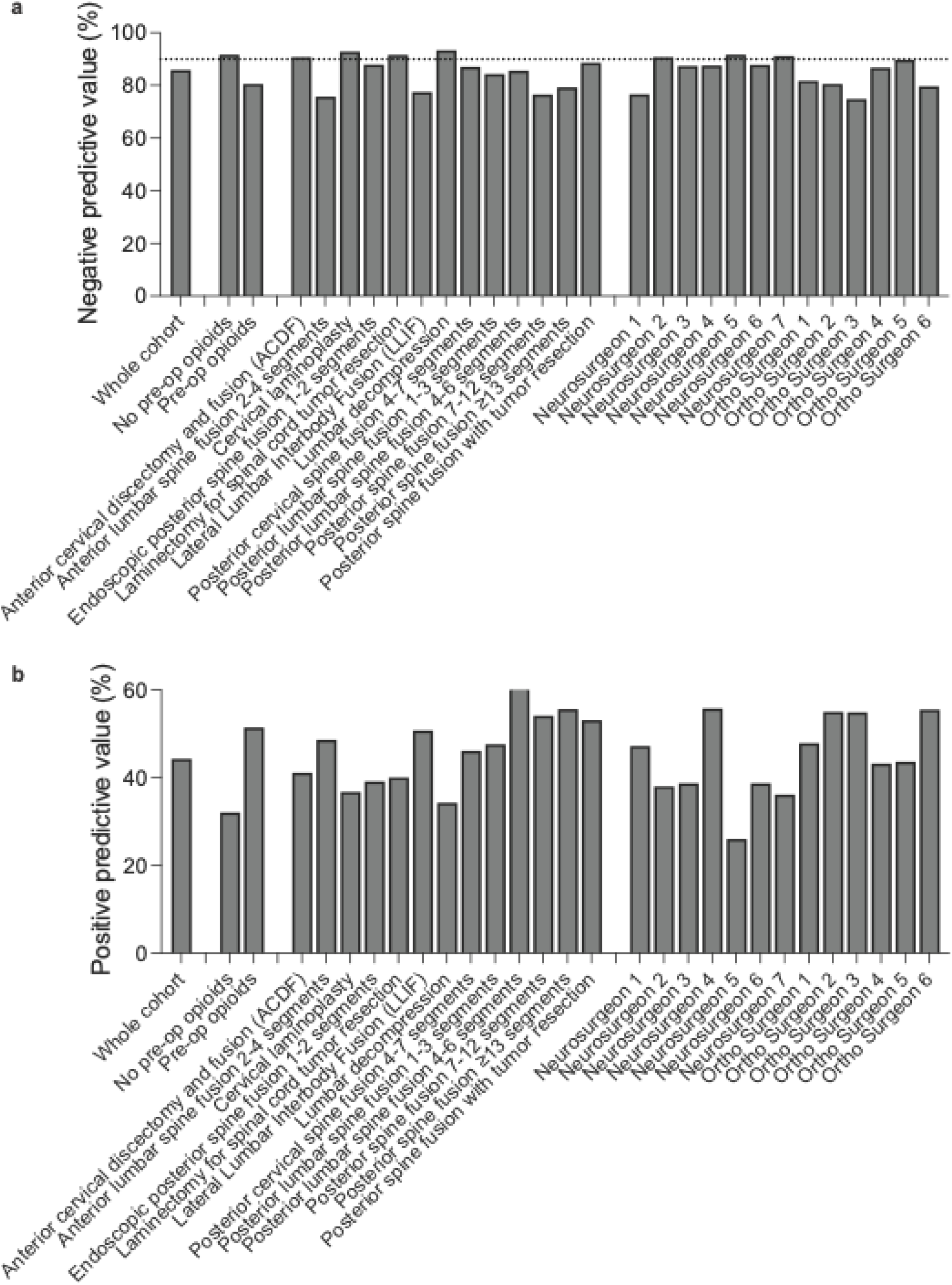
Using opioid refill at 1-30 days after discharge to predict a refill at 31-60 days. The negative (**a**) and positive (**b**) predictive values are presented for the whole cohort, with or without pre-operative opioids, across different procedures, and by different surgeons. Data were analyzed using two-sided Fisher’s exact test, with p<0.0001 for each analysis except neurosurgeon 5, which have p<0.01.

**Supplementary Table 1.**
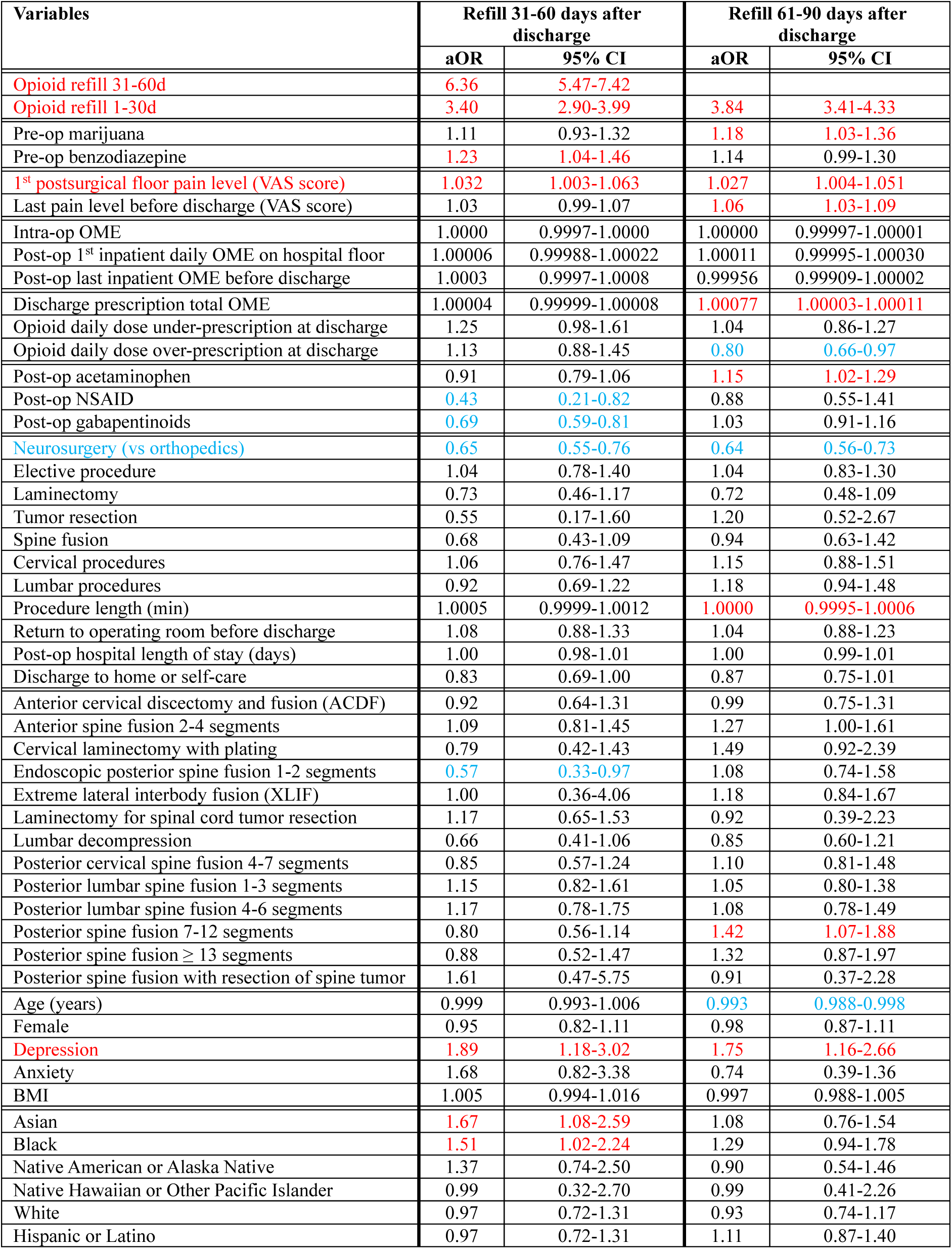
Independent risk factors associated with opioid refill prescriptions 61-90 and 31-60 days after discharge in patients who used opioids up to 6 months before their spine surgeries. aOR: adjusted odds ratio; CI: confidence interval.

**Supplementary Table 2.**
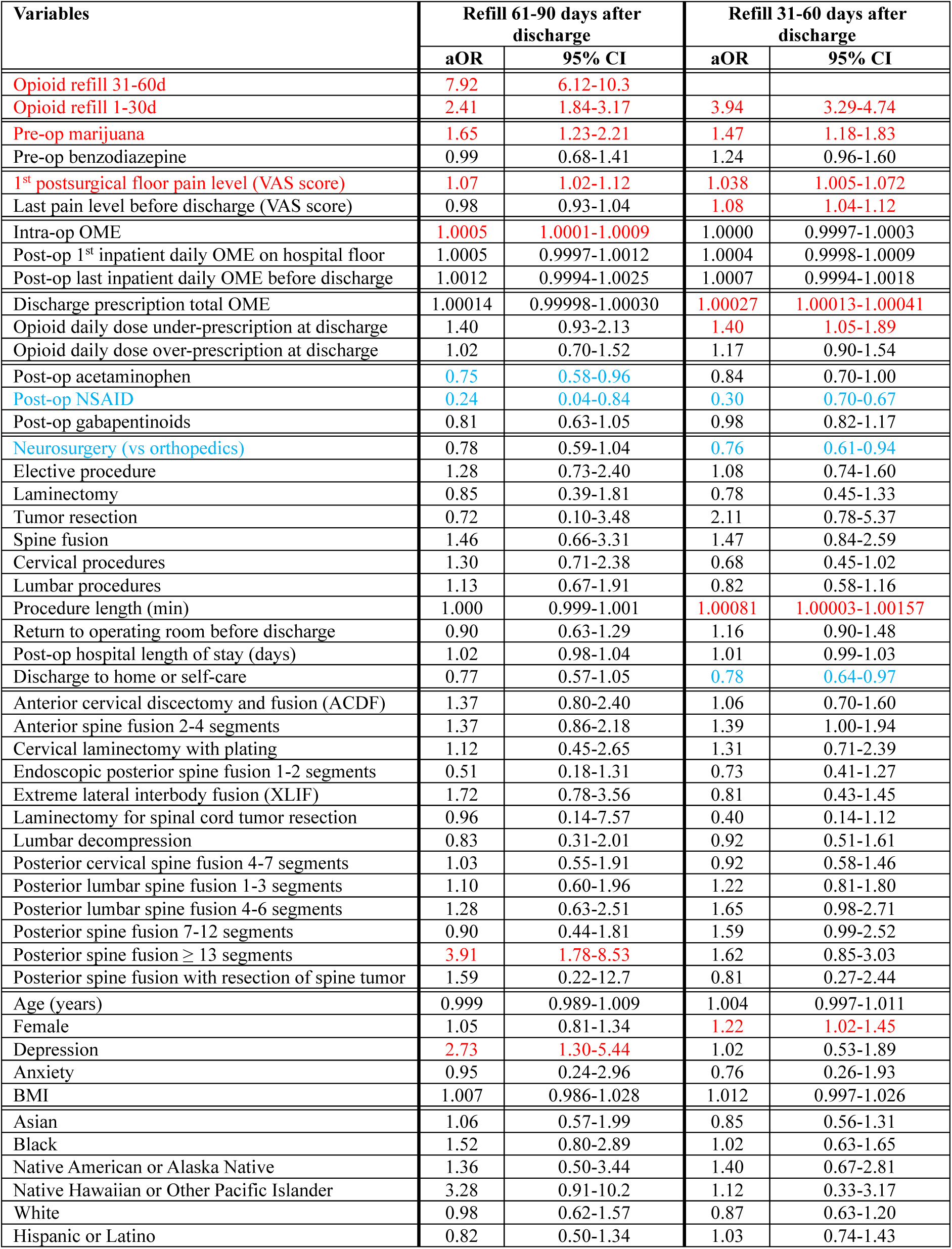
Independent risk factors associated with opioid refill prescriptions 61-90 and 31-60 days after discharge in patients who did not use opioids up to 6 months before their spine surgeries. aOR: adjusted odds ratio; CI: confidence interval.

**Supplementary Table 3.**
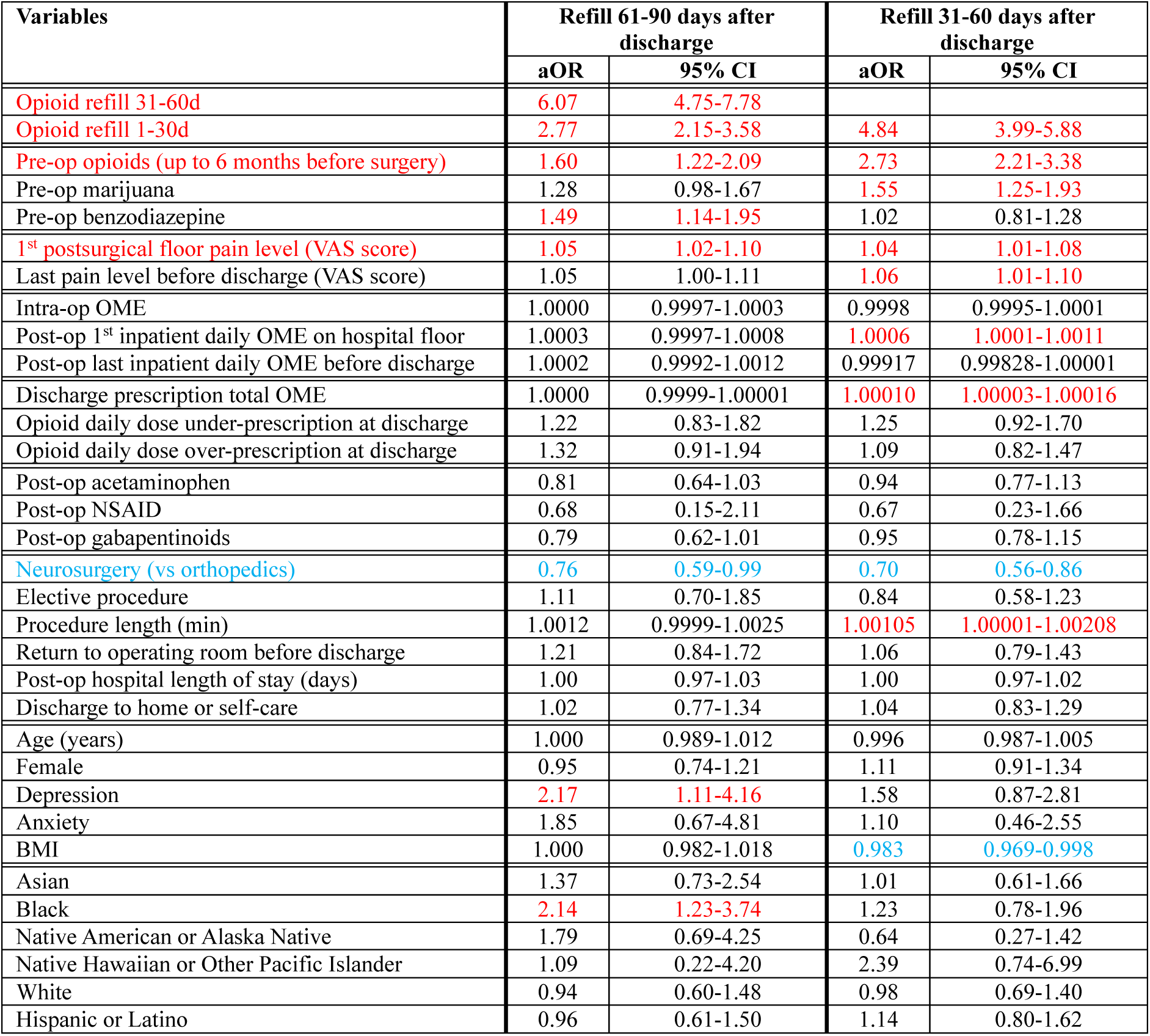
Independent risk factors associated with opioid refill prescriptions 61-90 and 31-60 days after discharge from cervical spine surgeries. aOR: adjusted odds ratio; CI: confidence interval.

**Supplementary Table 4.**
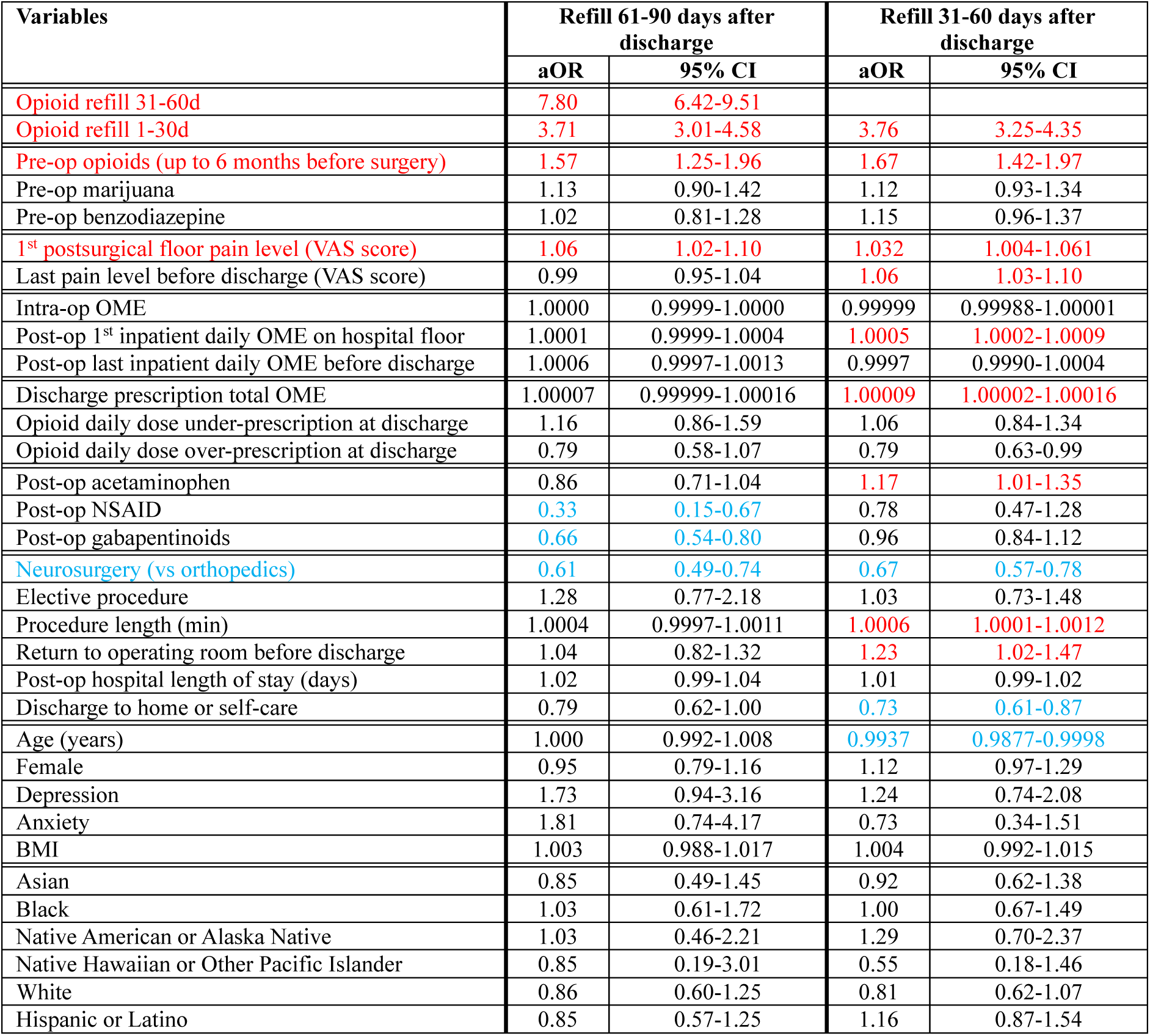
Independent risk factors associated with opioid refill prescriptions 61-90 and 31-60 days after discharge from lumbar spine surgeries. aOR: adjusted odds ratio; CI: confidence interval.

## Notes

### Competing Interest Statement

P.V.M. has served as a consultant for DePuy Spine, Globus, and Stryker; has direct stock ownership in Spinicity/ISD; has received royalties from DePuy Spine, Thieme Publishers, and Springer Publishing; and has received support from ISSG, NREF, NIH, and AOSpine for non-study-related clinical or research efforts.

### Funding Statement

This study did not receive any funding

### Author Declarations

IRB of the University of California San Francisco gave ethical approval for this work

### Summary of Updates

Editorial changes of the title and text, with no change in tables or figures

